# Measurement of Respiratory Rate using Wearable Devices and Applications to COVID-19 Detection

**DOI:** 10.1101/2021.05.15.21257200

**Authors:** Aravind Natarajan, Hao-Wei Su, Conor Heneghan, Leanna Blunt, Corey O’Connor, Logan Niehaus

## Abstract

We show that heart rate enabled wearable devices can be used to measure respiratory rate. Respiration modulates the heart rate creating excess power in the heart rate variability at a frequency equal to the respiratory rate, a phenomenon known as respiratory sinus arrhythmia. We isolate this component from the power spectral density of the heart beat interval time series, and show that the respiratory rate thus estimated is in good agreement with a validation dataset acquired from sleep studies (root mean squared error = 0.648 min^−1^, mean absolute percentage error = 3%). Using the same respiratory rate algorithm, we investigate population level characteristics by computing the respiratory rate from 10,000 individuals over a 14 day period, with equal number of males and females ranging in age from 20 - 69 years. 90% of respiratory rate values for healthy adults fall within the range 11.8 min^−1^ 19.2 min^−1^ with a mean value of 15.4 min^−1^. Respiratory rate is shown to increase with nocturnal heart rate. It also varies with BMI, reaching a minimum at 25 kg/m^2^, and increasing for lower and higher BMI. The respiratory rate decreases slightly with age and is higher in females compared to males for age < 50 years, with no difference between females and males thereafter. The 90% range for the coefficient of variation in a 14 day period for females (males) varies from 2.3%−9.2% (2.3%−9.5%) for ages 20−24 yr, to 2.5%−16.8% (2.7%−21.7%) for ages 65−69 yr. We show that respiratory rate is often elevated in subjects diagnosed with COVID-19. In a 7 day window centered on the date when symptoms present (or the test date for asymptomatic cases), we find that 33% (18%) of symptomatic (asymptomatic) individuals had at least one measurement of respiratory rate 3 min^−1^ higher than the regular rate.

## I. INTRODUCTION

It is well known that heart rate varies with respiration, increasing during inhalation, and decreasing during exhalation. This modulation of the heart rate in response to respiration is known as Respiratory Sinus Arrhythmia (RSA), and is associated with the efficiency of pulmonary gas exchange [1–3]. RSA thus manifests as excess power at the respiration frequency, making it possible to infer the respiratory rate from heart beat interval data.

Unlike other vital signs such as pulse rate and blood pressure, the respiratory rate can be consciously altered by a patient who is aware of the measurement being made, potentially resulting in flawed recordings. The respiratory rate is a valuable metric in determining clinical deterioration [4, 5] and an increase of 3 min^−1^ to 5 min^−1^ can indicate deterioration [4]. The heart rate to respiratory rate ratio and respiratory rate to oxygen saturation ratio have been shown to be useful indicators in predicting the duration of hospitalization [6]. In a study of patients admitted to the hospital with pneumonia from 2010 - 2012, it was shown that those with a respiratory rate in excess of 27 min^−1^ had an odds ratio of 1.72 for in-hospital death [7]. The respiratory rate factors into the CURB-65 score for predicting mortality in community-acquired pneumonia [8], as well as during epidemics [9]. Elevated respiratory rate values (> 27 min^−1^) have been shown to be predictive of cardiopulmonary arrest [10]. Increased respiratory rate factors into early warning scores meant to assess the likelihood of a patient needing critical care [11–13]. The respiratory rate has also been shown to be a useful biomarker for COVID-19 detection [14, 15]. Despite these findings, the respiratory rate is not always recorded while monitoring patients, and may be considered a neglected vital sign [6, 16, 17].

The clinical value in measuring respiratory rate, and the growing interest in wearable devices provides a valuable opportunity in the field of digital health. Wearable devices can compute the respiratory rate during sleep, thus obtaining measurements that are made without the conscious knowledge of the user. Commercial wearable devices accomplish this through photoplethysmography (PPG) [18–20], usually at a single point of contact, either on the wrist (smartwatches, trackers, straps) or the finger (rings). Respiration modifies the PPG time series signal in a number of ways [21–23]. In this work, we focus on the RSA feature, i.e. the frequency modulation of the PPG.

Karlen et al. [22] computed the respiratory rate from PPG in a study involving both children and adults, and found agreement with capnometry measurements up to respiratory rates of ∼ 45 min^−1^. In a study involving 32 participants, it was shown that the respiratory rate computed by WHOOP devices compared well with polysomnography (PSG) measurements [24], with low bias (1.8%) and precision error (6.7%). The respiratory rate may also be inferred from the PPG time series through deep learning techniques [25]. In the present work, we briefly describe how the respiratory rate may be inferred from the RSA feature in the power spectral density of heart beat interval time series data. We compute the respiratory rate over the course of a night, and show that it agrees well with validation data obtained from ground truth measurements. We examine how the respiratory rate varies with age and sex, and how much it varies relative to the mean value over the course of 14 days. We also investigate its dependence on BMI and heart rate. Finally, we build upon earlier work [15] and show that the respiratory rate can be a valuable biometric in the detection and monitoring of COVID-19.

## II. MATERIALS AND METHODS

### A. Data

#### 1. Respiratory rate data

The dataset used to explore correlations between respiratory rate and age, sex, BMI, and heart rate consisted of 10,000 users of Fitbit devices who reside in the United States or Canada, and who wore their devices to sleep in the date range Nov 1 − 14, 2020. We collected sleep and heart rate variability data from these Fitbit users during this 14 day period. The data were collected and anonymized consistent with Fitbit’s terms and conditions. The dataset consisted of male and female individuals in the age range 20-69 with 500 subjects of each sex and each of 10 equally spaced age bins (5 year age bin size), yielding a total of 135,947 usable measurements. The mean Body Mass Index (BMI) of the participants was 27.8 ± 5.2 for males and 27.5 ± 6.4 for females, where the quoted error bar is 1 standard deviation. The main Fitbit devices used to collect these data include Charge 3 (22.5%), Versa 2 (20.0%), Inspire HR (11.3%), Versa (10.0%), Charge 2 (9.62%), and Charge 4 (7.68%), with a number of other devices contributing less than 5% each.

#### 2. Validation data

We conducted 2 experiments to validate the respiratory rate algorithm. Experiment A was conducted at Sleep Med in Columbia, SC, from Oct 17, 2019 to Nov 6, 2019, and used a polysomnography device (Alice 5). Experiment B was conducted remotely, by shipping equipment to the homes of participants, from March 9, 2020 to May 29, 2020, and used a Home Sleep Test (Resmed Apnealink). Both experiments were approved by an Institutional Review Board (Solutions IRB). Participants provided informed consent for their data to be collected and used for research purposes. Participants in Experiment A wore Fitbit devices on both wrists, while participants in Experiment B wore a Fitbit device on one wrist only. We excluded participants with severe sleep apnea (Apnea-Hypopnea Index ≥ 30). 52 measurements were obtained from 28 individuals (15 female, 13 male) between the ages of 32 and 71 (mean age was 48.9 yr with a standard deviation of 9.5 yr). More details regarding the data collection may be found in Table S1 in the Supplementary Text.

#### 3. COVID-19 data

The Fitbit COVID-19 survey is an ongoing survey of Fitbit users residing in the United States or Canada. Participants provided information on whether they were diagnosed with COVID-19, and whether they experienced symptoms. The data for the COVID-19 survey were collected with Institutional Review Board approval (Advarra IRB), and participants provided written consent for their data to used for research purposes. The data used in the present study comprises a subset, consisting of 3,236 individuals with COVID-19 PCR positive test dates (self reported) ranging from Feb 28 - Nov 13, 2020, with 2,939 symptomatic and 297 asymptomatic individuals. 77.6% of participants identified as female. The mean age was 42.25 ± 12.35 yr, and the mean BMI was 30.29 ± 7.25, where the stated errors are 1 standard deviation. More details regarding the Fitbit COVID-19 survey may be found in Ref.[15].

#### 4. Software

All statistical analyses were performed using standard Python packages such as NUMPY and SCIPY. The respiratory rate code software was written in Scala and uses the BREEZE library.

### B. Computation of respiratory rate from heart rate variability

Interbeat interval values are computed from the heart beat interval time series data and assembled into non-overlapping 5 minute blocks. The data are cleaned to remove noise due to motion artifacts, electronic artifacts, missed heart beats, etc. For details on the cleaning and pre-processing steps, we refer the reader to Ref.[26]. Each 5 minute block of data is resampled to obtain 512 equally spaced samples allowing us to resolve all frequency components up to 0.5 × (512*/*300) = 0.85 Hz. The resolution in frequency space is 1/300 Hz. The mean of the data in the time window is subtracted, and the data smoothed with a Hann window. A Fast Fourier Transform is applied, and properly normalized to give us the Power Spectral Density (PSD), which is the power contained per unit frequency. Integrating the PSD over the range 0.04 Hz - 0.15 Hz gives us the low frequency (LF) power, while integrating the PSD over the range 0.15 Hz - 0.4 Hz gives us the high frequency (HF) power. The PSD for different 5 minute segments are aggregated.

The PSD of HRV fluctuations is shown in Fig. 1 for a single individual and for one night: The plot contains 2 main components: *background* and *RSA*. To isolate the RSA component, we need to model the background and subtract it from the power spectrum.

**FIG. 1.**
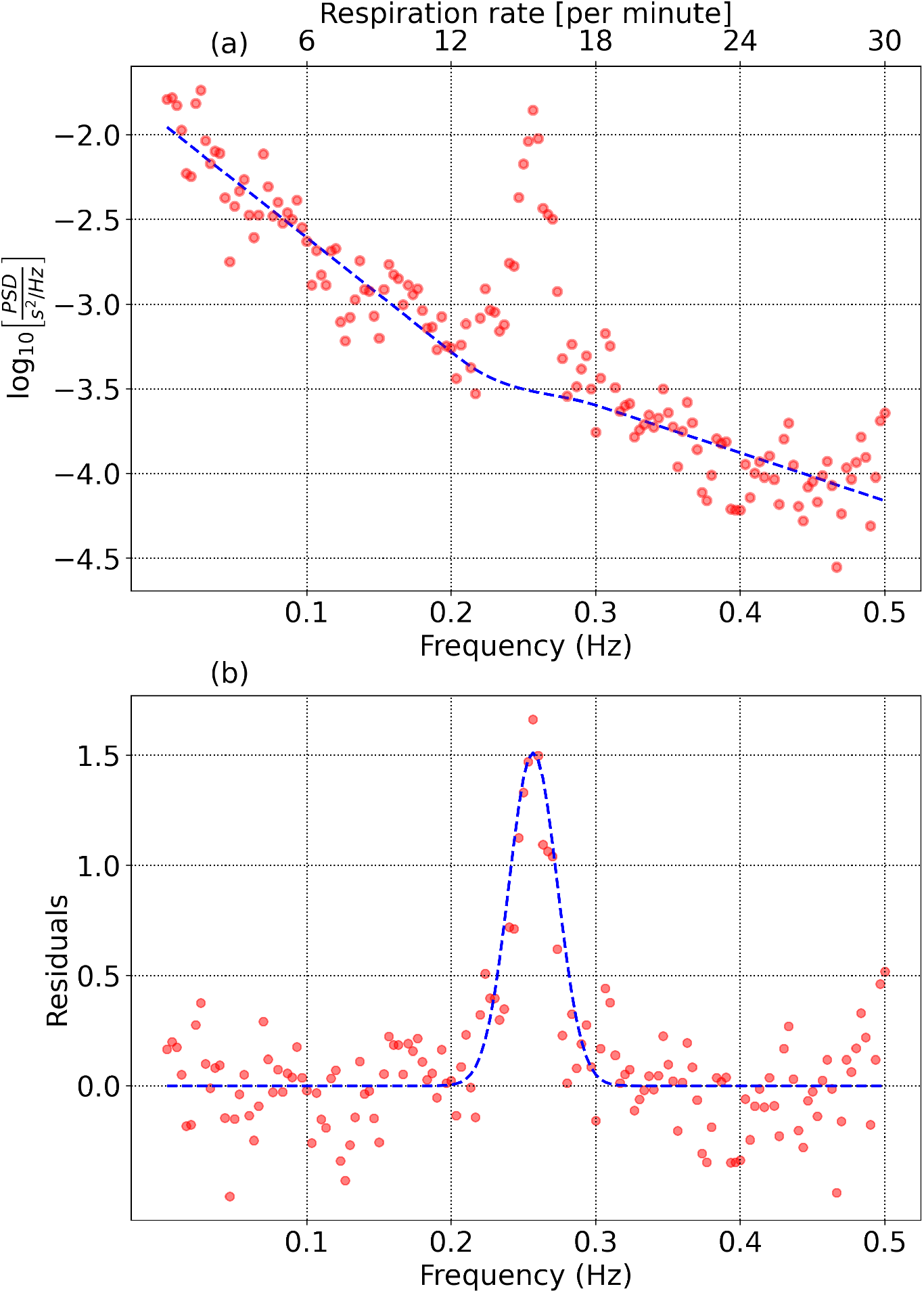
(a) HRV power spectral density consisting of background and Respiratory Sinus Arrhythmia. (b) residuals after the background is subtracted.

#### 1. Modeling the background

We set a maximum frequency *f*_max_ = 0.5 Hz (corresponding to a respiratory rate of 30 min^−1^), and discard data at higher frequencies. We also set a minimum frequency *f*_min_= 0.1367 Hz (corresponding to a respiratory rate of 8.2 min^−1^). The power spectrum at frequencies from *f*_0_ = 1*/*300 Hz to *f*_min_ is used to determine the noise level. The RSA feature is contained between two frequencies *f*_1_(> *f*_min_) and *f*_2_(< *f*_max_) which we will determine iteratively.

1. Low frequency background: The PSD from frequencies *f*_0_ to *f*_1_ is modeled by a function of the form log_10_[PSD] = *c*_1_ + *c*_2_*f*.
2. High frequency background: The PSD from frequencies *f*_2_ to *f*_max_ is modeled by a similar function: log_10_[PSD] = *c*_3_ + *c*_4_*f*.
3. The PSD from *f*_1_ to *f*_2_ is modeled by a *patching function*: log_10_[PSD] = *p*_1_ + *p*_2_*f* + *p*_3_*f* ^2^ + *p*_4_*f* ^3^. *p*_1_ and *p*_2_ are fixed to match the end points of the low frequency and high frequency background regions, while *p*_3_ and *p*_4_ are set to match the derivatives at the end points, thus enabling a smooth transition.

#### 2. Isolating the signal

To begin, we assign reasonable values to *f*_1_ and *f*_2_, which will be refined in subsequent iterations. We initialize *f*_1_ = *f*_min_ and *f*_2_ = 0.333 Hz (corresponding to a respiratory rate of 20 min^−1^). In practice, the choice of *f*_1_ and *f*_2_ are determined by the expected range of respiratory rates in the population under study. Signal estimation is performed using the following steps:

1. The power spectrum is modeled as described earlier, and parameterized by the variables (*c*_1_, *c*_2_, *c*_3_, *c*_4_, *p*_1_, *p*_2_, *p*_3_, *p*_4_).
2. The background function is subtracted from the data to obtain the residuals. The residuals are low pass filtered (we use a median filter of size 3) to reduce noise, and interpolated (we use a cubic spline) to maintain the original frequency resolution.
3. The peak of the residuals is identified as *A*_RSA_, and the frequency corresponding to the maximum value = *f*_RSA_. Assuming a gaussian distribution for the RSA feature, we identify a frequency *f*_−_ < *f*_RSA_ such that *A*(*f*_−_) = 0.6065*A*_RSA_, as well as a frequency *f*_+_ > *f*_RSA_ such that *A*(*f*_+_) = 0.6065*A*_RSA_. The mean of these two values *f*_resp_ = 0.5 × (*f*_+_ + *f*_−_) is identified as the mean respiratory frequency. The standard deviation is *σ* _resp_ = 0.5 × (*f*_+_ *−f*_−_). The mean *µ*_noise_ and standard deviation *σ*_noise_ of the residuals from *f*_0_ to *f*_min_ are calculated. The signal−to-noise ratio *SNR* is defined as *SNR* = (*A*_RSA_ *-µ*_noise_) */σ*_noise_.
4. *f*_1_ is redefined as *f*_resp_ − 3*σ*_resp_, and *f*_2_ is set to *f*_resp_ + 3*σ*_resp_.

Steps 1-4 are repeated until either successive estimates of *f*_resp_ agree to within 1%, or 5 iterations are completed. We restrict our range of respiratory rates to between 10 min^−1^ to 26 min^−1^. Frequencies much higher than 26 min^−1^ are hard to resolve due to the rapid fall-off of the power spectral density with frequency, while resonances at frequencies lower than 10 min^−1^ may be confused with Mayer wave oscillations [27]. The values of (*f*_resp_, *σ*_resp_, *SNR*) are stored for each individual, for each day, provided SNR ≥2.5. Fig. 1(b) shows the residuals and estimation of the RSA feature. Also shown is a gaussian with mean *f*_resp_ and standard deviation *σ*_resp_.

When aggregating respiratory rate measurements over multiple days, we adopt a numerical approach: The respiratory rate measurement for any given day for each individual is treated as a random variable drawn from a gaussian distribution with mean *f*_resp_ and standard deviation *σ*_resp_. We randomly choose 100 samples from this distribution for each day. The mean and standard deviation over all samples is then computed. We follow the same process for averages involving multiple subjects.

### C. Validation of estimated respiratory rate data with ground truth measurements

We obtained 52 measurements of air flow data, from 28 individuals through polysomnography (PSG), or a home sleep test (HST). Data were collected from 1 to 3 nights for each participant, with devices on either one or both wrists (data from the two experiments were combined, see Table S1 in the Supplementary Text for details). Data from the air flow sensor were band pass filtered with a fourth order Butterworth filter to retain frequencies between 10 min^−1^ - 30 min^−1^. The data were then analyzed with the help of a spectral peak detection algorithm with a window size of 51.2 s and a step size of 6.4 s. The median of all respiratory rate measurements over the night is computed, and serves as the true respiratory rate.

Fig. 2 shows the comparison between the true respiratory rate and the rate estimated from the peak of the heart beat interval power spectral density. Plot (a) shows 52 measurements in the range (10 min^−1^, 26 min^−1^) with SNR ≥2.5, obtained from 28 individuals with apnea-hypopnea index < 30. The Pearson correlation coefficient *r* = 0.9515. Plot (b) shows the Bland-Altman plot of the difference in measurements (predicted value - true value) plotted against the average of the two. The bias (mean of the difference between predicted and true values) is 0.244 min^−1^ (1.67%), and the root mean squared error is 0.648 min^−1^ (4.2%). The mean absolute error is found to be 0.460 min^−1^, and the mean absolute percentage error = 3.0%.

**FIG. 2.**
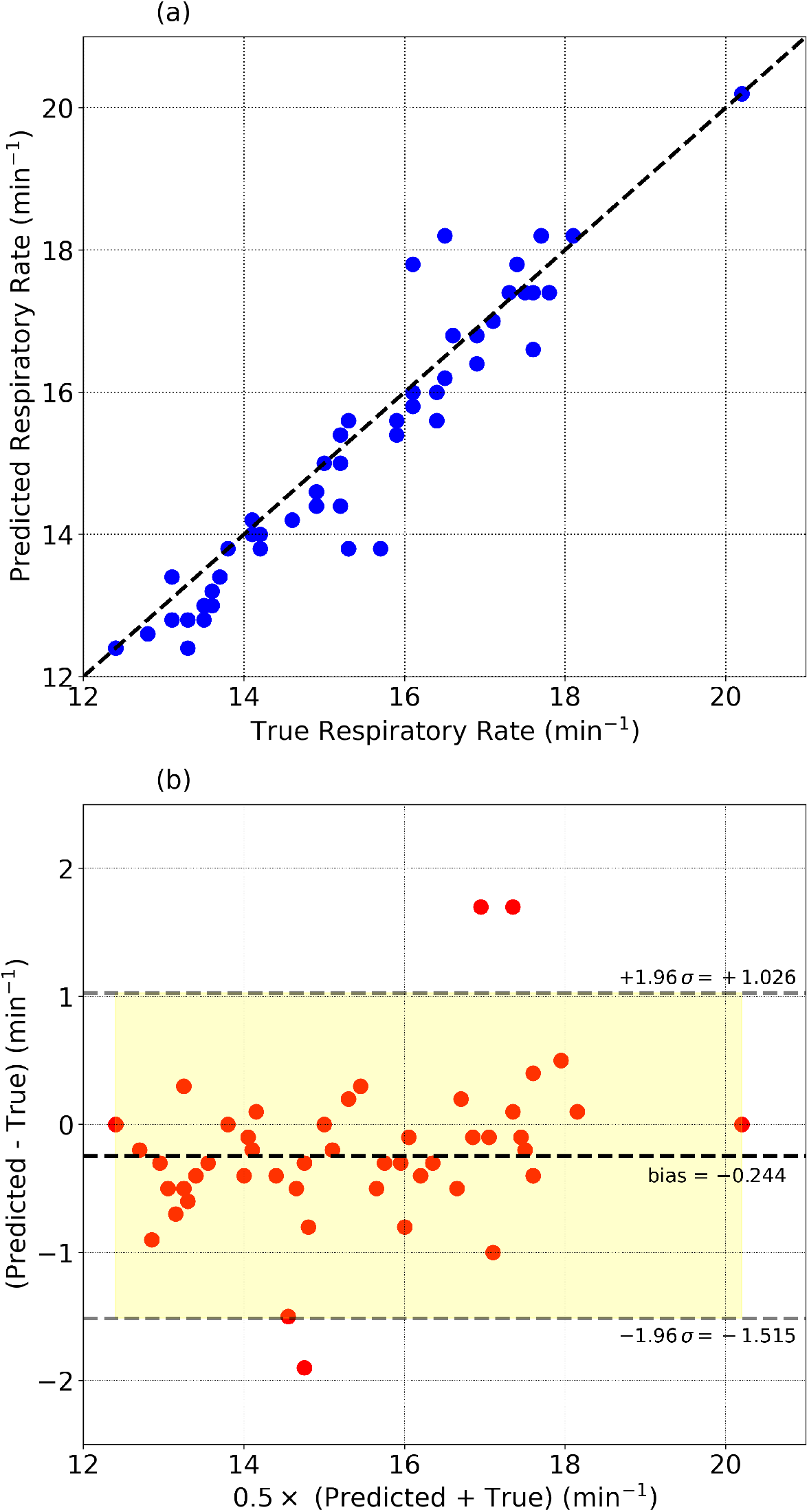
(a) Respiratory rate estimated from the heart beat interval time series data compared to ground truth measurements. (b) shows the Bland-Altman plot comparing the true and predicted values. The bias (mean of predicted value - true value) is -0.24 min^−1^ (−1.67%). The RMS error = 0.65 min^−1^ and the mean absolute error = 0.46 min^−1^ (3.0%). The 95% region is shown in yellow.

## III. RESULTS

Sleep consists of 3 main stages: Light sleep (stages *N*_1_ and *N*_2_), deep sleep (stage *N*_3_), and REM sleep. Fitbit has developed a validated algorithm which estimates a person’s different sleep stages over a night. Fitbit heart rate and sleep measurements have been studied by an external group who found that Fitbit Charge HR devices showed a 97% sensitivity and a 91% accuracy in detecting sleep [28]. It is known that different stages of sleep are likely to have varying magnitudes of respiratory sinus arrhythmia [29]. The sinus arrhythmia component is contained within the HF band for respiratory rate values > 9 min^−1^. Thus the HF power can serve as a proxy for the magnitude of sinus arrhythmia. Let us define the dimensionless metric HF_*v*_ = HF / (HF + LF). The value of HF_ν_ averaged over all individuals in deep sleep is found to be HF_*v*,deep_ = 0.40 ± 0.17 (stated values are mean and standard deviation). In Light sleep, the equivalent HF_*v*,light_ = 0.27 ± 0.13, while in REM sleep, we find HF_*v*,REM_ = 0.19 ± 0.11. We thus find with our data that HF power is largest in deep sleep, and least during REM sleep. For the following results, we ignore REM sleep, and estimate the respiratory rate primarily during deep sleep if SNR_deep_ ≥2.5 is obtained and during light sleep (provided SNR_light_ ≥2.5) if SNR_deep_ < 2.5. We note that in the validation test described in the Materials and Methods section, we computed respiratory rate during all sleep stages since we did not have sleep stage information for the data collected with the PSG and HST. A large difference in respiratory rate between sleep stages is not expected according to Ref. [30–32]. However, Ref. [33] found a statistically significant increase from 16.1 ± 2.0 min^−1^ in non-REM sleep to 17.9 ± 2.7 min^−1^ in REM sleep (*p* < 0.05). Ref.[34] also found a statistically significant difference in respiratory rate among sleep stages (*p*< 0.001), with REM sleep having the highest rate (*p*< 0.01).

We estimated the probability of the algorithm taking 0,1,2,3,4, and 5 iterations to estimate the respiratory rate, using a subset of 1,000 randomly selected individuals on one night of data (0 iterations means there was either no data, or the signal-to-noise ratio was found to be too low for a reliable estimate. 14.6% of measurements had 0 iterations, i.e. no result with deep sleep data, 6.1% of measurements had no result with light sleep data, and 2.6% of measurements had no result with either deep or light sleep data). For measurements in deep sleep, the fraction of estimates taking 1,2,3,4,5 iterations were respectively, 50.8%, 22.6%, 4.2%, 1.3%, and 6.5%. For measurements in light sleep, the fractions were found to be 50.7%, 28.6%, 5.7%, 1.2%, and 7.7%. These results assume a convergence threshold of 1% between successive iterations. Note that respiratory rate estimates that take 5 iterations may not have attained the required level of convergence (if the convergence threshold is relaxed to 5%, only 1% of measurements in deep sleep and 0.9% in light sleep required 5 iterations).

Fig.3 shows the distribution of respiratory rate values, with a bin size of 1 min^−1^. 90% of values fall in the range 11.8 min^−1^ − 19.2 min^−1^. The 95% range is 11.2 min^−1^ − 20.0 min^−1^. The mean of the distribution is 15.4 min^−1^ and the standard deviation is 2.35 min^−1^.

**FIG. 3.**
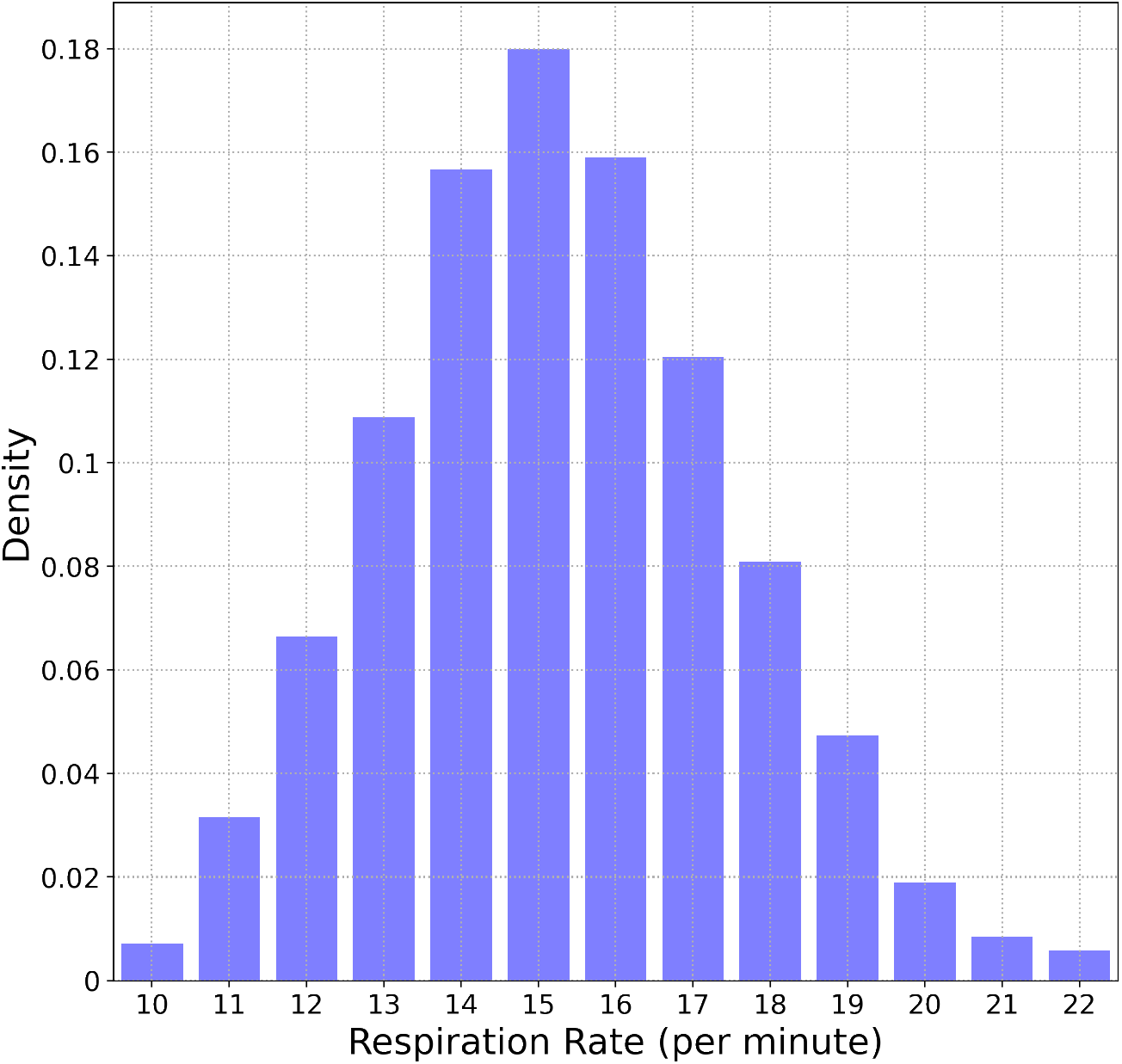
Distribution of respiratory rates. 90% of values are between 11.8 - 19.2 min^−1^.

Fig. 4(a) shows the variation of respiratory rate with age and sex. The black points show the measurements for female participants, while the green dots represent males. The age bin size is 5 years, and the error bars are one standard deviation. The respiratory rate for females is higher than for males for age < 50 yr (*p −* value < 0.001). There is no statistically significant difference between males and females for age > 50 yr. The mean respiratory rate for females (males) decreases from 16.7 (15.5) min^−1^ in the age group 20 yr - 24 yr, to 14.8 (14.8) min^−1^ in the age group 65 yr - 69 yr, a difference of 1.9 (0.7) min^−1^ for females (males) over a span of 50 yr. For age below 50 yr, the Pearson *r* correlation coefficient comparing the dependence of mean respiratory rate with age for females (males) is − 0.145(−0.104). For ages > 50 yr, we find *r* = − 0.031(−0.043) for females (males).

**FIG. 4.**
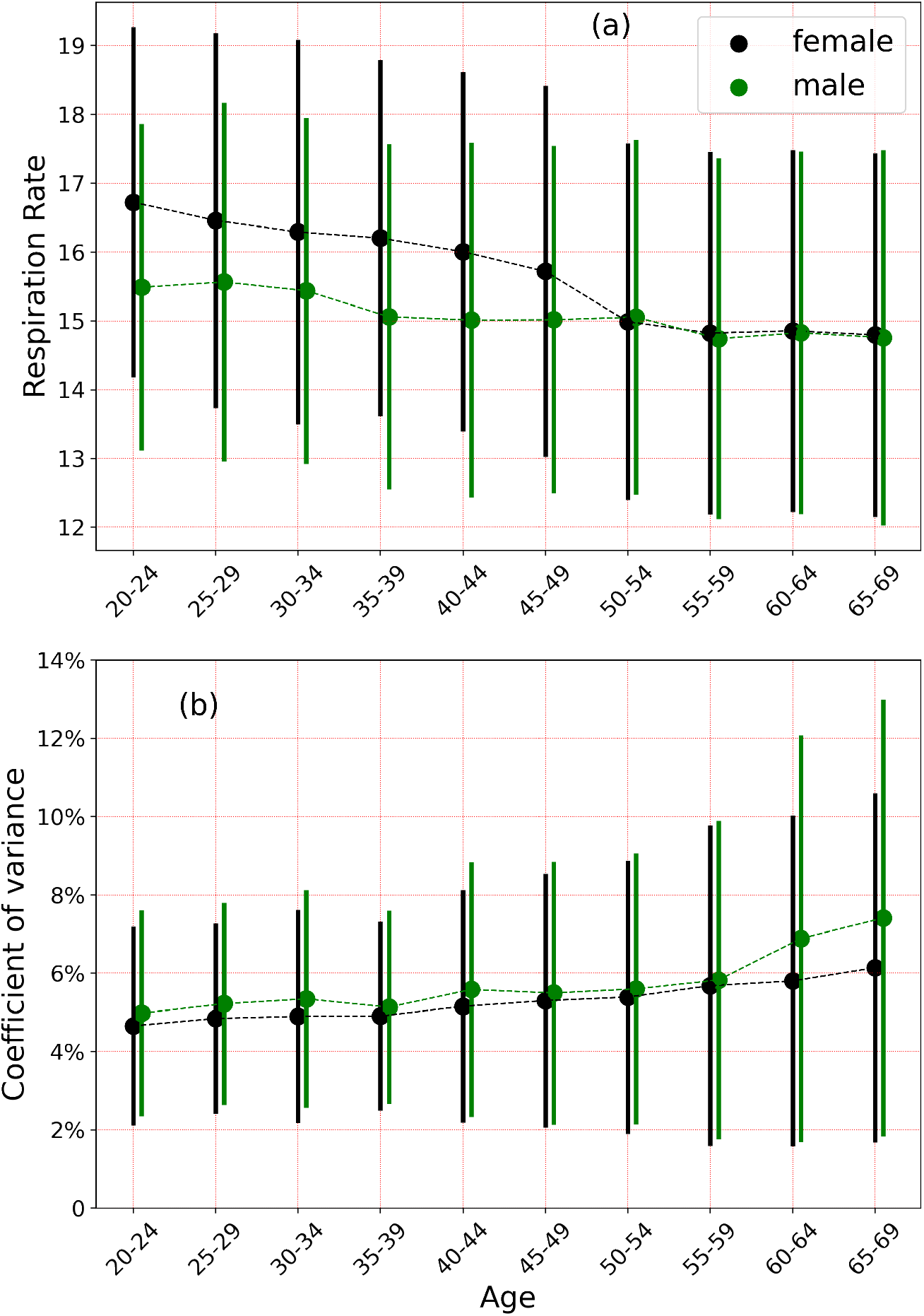
(a) shows the variation of respiratory rate with age and sex. Females have a higher respiratory rate on average for ages < 50 yr, and no difference thereafter. (b) shows the coefficient of variation over a 14 day period. Error bars are 1 standard deviation.

Plot (b) shows the coefficient of variance (CoV) (ratio of standard deviation to the mean) measured over a 14 day period, and only considering subjects with 10 or more nights of data. The CoV increases with age, with a Pearson *r* −correlation coefficient of 0.132 (0.172) for females (males). The CoV varies from 4.65% (4.98%) in the age range 20 yr - 25 yr to 6.14% (7.41%) in the age range 65 yr - 69 yr for females (males). The difference between male and female participants is most significant above age 60 yr (*p* − value < 0.001).

The dependence of respiratory rate with BMI (measured in kg/m^2^) is shown in Fig. 5(a). The bin size in BMI = 1 and the error bars represent the standard error of the mean. The respiratory rate reaches a minimum at BMI ≈ 25. For BMI < 25, the respiratory rate decreases with increase in BMI, with a Pearson *r* −correlation coefficient of -0.0425. For values of BMI ≥25, we see an increase with BMI, with *r* = 0.196. Expanding in a Taylor series about the minimum, we find that the mean respiratory rate *R* measured in min^−1^ may be expressed as:

**FIG. 5.**
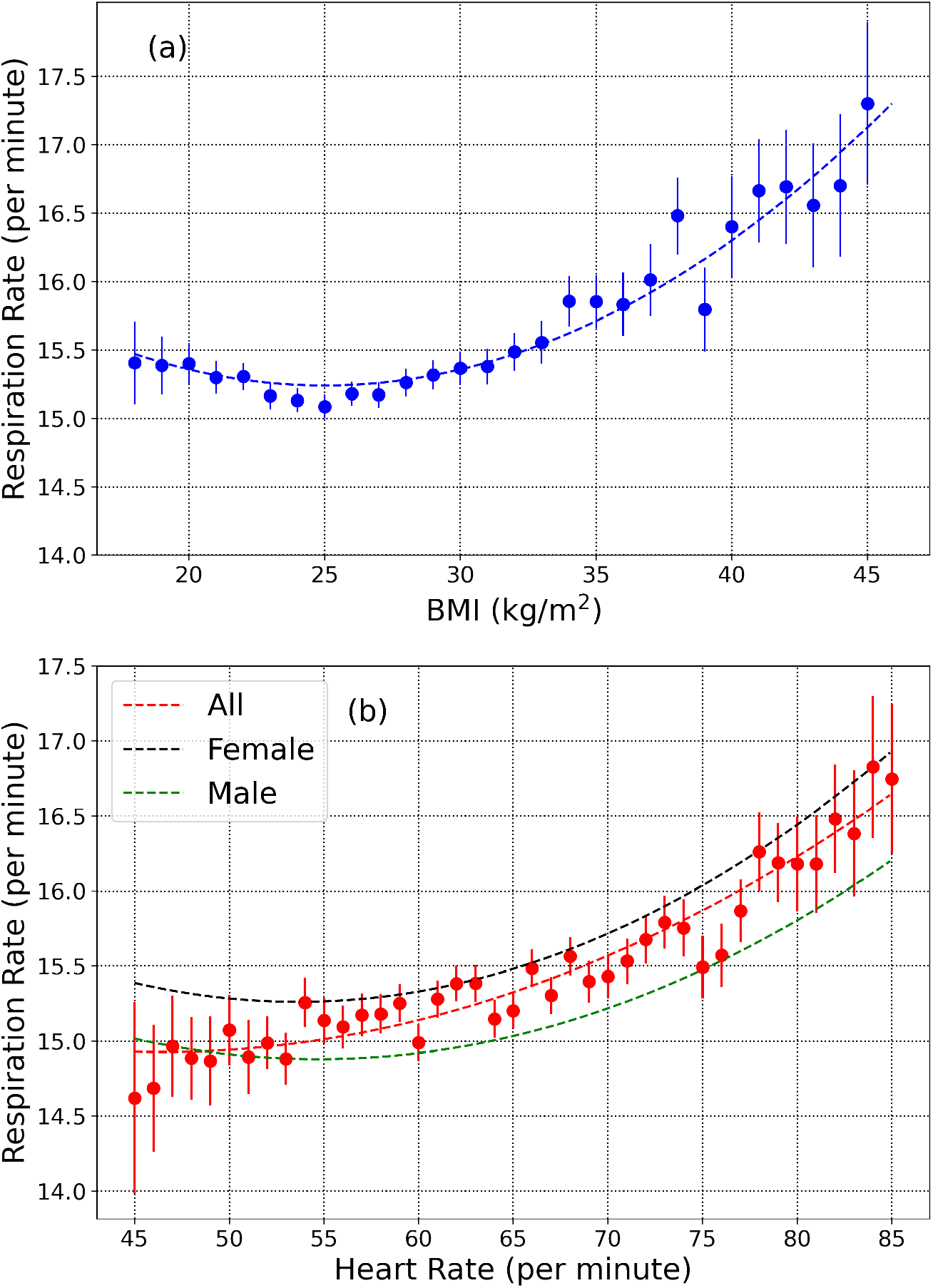
(a) respiratory rate dependence on BMI. The lowest value occurs at a BMI of ∼ 25. (b) respiratory rate variation with nocturnal heart rate measured in non-REM sleep (black and green curves are for females and males, respectively, the red curve is for all participants). Error bars show the standard error of the mean.

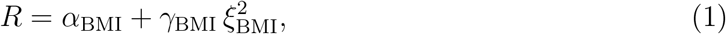

where *α*_BMI_ = 15.24, *γ* _BMI_=2.95. 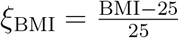. Eq.1 is a useful model over the range of BMI 18−45.

The variation of respiratory rate with nocturnal heart rate is shown in Fig. 5(b). The heart rate in beats per minute (bpm) is measured in non-REM sleep. The red curve is for all individuals, while the black and green curves are plotted for female and male participants respectively. The mean respiratory rate (for all participants) increases with increase in heart rate, with a Pearson *r* –correlation of 0.154. It is possible to model the mean respiratory rate *R* (measured in min^−1^) dependence on heart rate as:

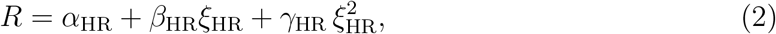

where *α*_HR_ = 15.14,, *β*_HR_ = 1.88, *γ*_HR_ = 4.17. 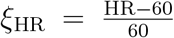. Eq.2 was fitted for all participants (male and female), and is useful over the range 45 85 bpm.

### A. Effect of COVID-19 on the respiratory rate

In this section, we present results from a subset of the Fitbit COVID-19 data survey. Let *µ* and *σ* be the mean and standard deviation of the respiratory rate for a specific user, estimated several days prior to the onset of illness. The *Z* –score on a given day *D*_*n*_ may be defined as

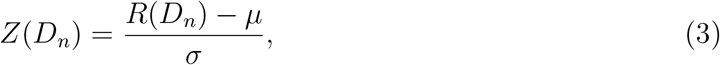

where *R*(*D*_*n*_) is the respiratory rate for a specific user on day *D*_*n*_. For symptomatic individuals, let *D*_0_ be the date when symptoms present. For asymptomatic users, we set *D*_0_ to the test date. Mean and standard deviation of the respiratory rate are computed using data from *D*_−90_ − *D*_−30_, only considering users with at least 30 days of data in this date range. There were 1,247 symptomatic individuals (from a total of 2,939) and 133 asymptomatic individuals (from a total of 297) satisfying this requirement. Fig. 6(a) shows the average *Z* –score measured for symptomatic individuals. The *Z* –score ≈ 0 for days < *D*_−14_, but increases thereafter, reaching a peak on *D*_+2_, i.e. two days following the day when symptoms first present. Interestingly *Z* does not fall off to zero, but instead approaches a constant between *D*_+14_ − *D*_+28_.

**FIG. 6.**
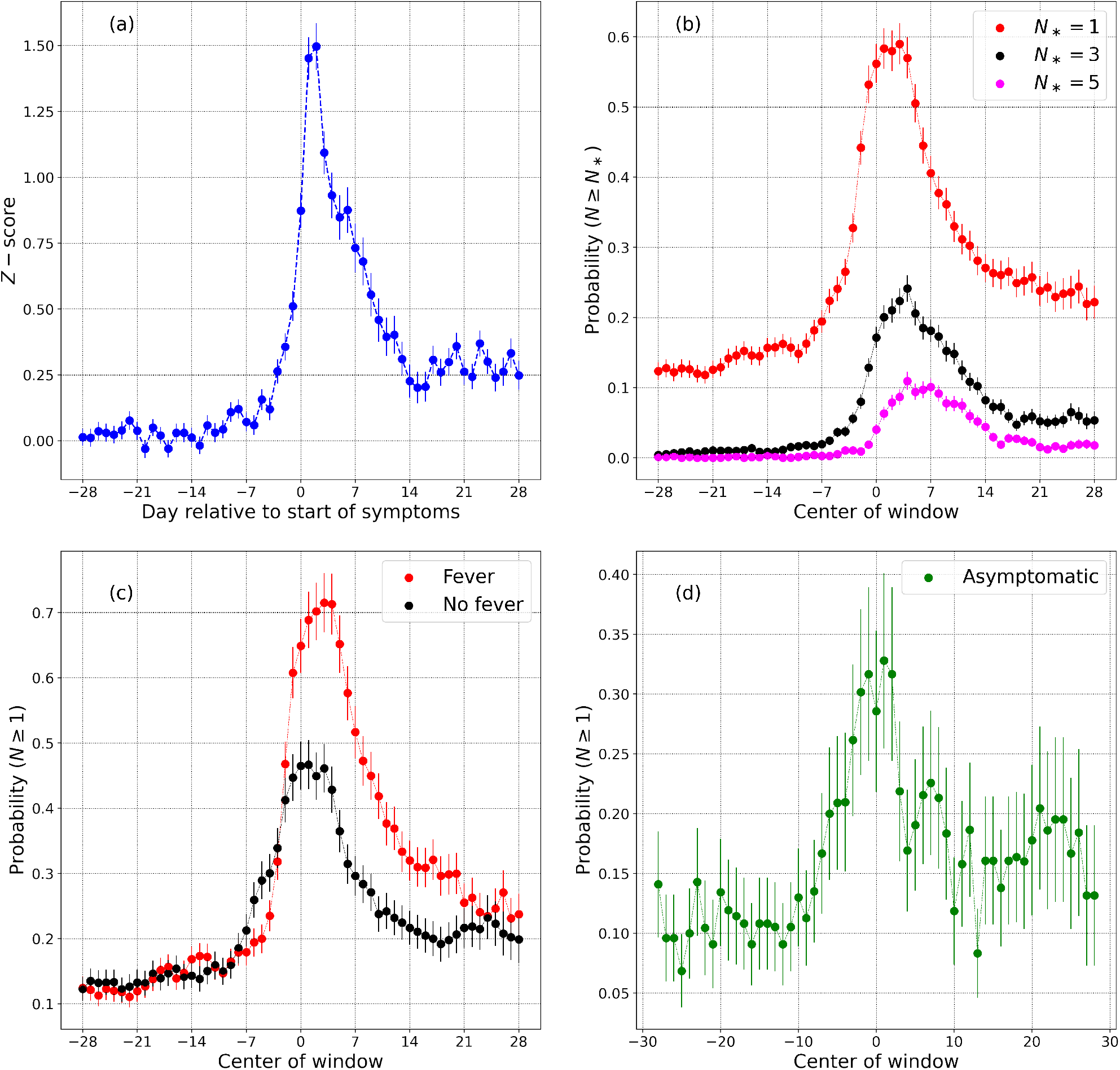
(a) shows the dimensionless *Z* –scored respiratory rate in symptomatic individuals, with day relative to the start of symptoms (Day 0 is the day when symptoms present). (b) measures the probability of receiving *N ≥ N*_*\**_ anomalously high values in a 7 day window centered on day *D*, for *N*_*\**_ = 1, 3, 5. The effect of fever is seen in (c). The variation of respiratory rate for asymptomatic individuals is shown in (d). Error bars show the standard error of the mean.

Next, we investigate the likelihood that a randomly selected symptomatic individual will receive an anomalously high respiratory rate value on a specific day. Let us consider a 7 day window, and compute the probability that a subject with receive *N* ≥*N*_*_ respiratory rate values satisfying *Z* ≥2.326 (this threshold corresponds to a *p –*value of 0.01 for a 1-tailed test. We are only concerned with values above the mean). Plot (b) shows the results for 7-day windows centered from *D*_−28_ to *D*_+28_, only considering subjects with all 7 days of valid data in the window. Shown are probabilities for *N*_*_ = 1,3, and 5. Plot (c) shows the effect of fever which is known to increase the respiratory rate [35]. The red data points show the probability for *N*_*_ = 1 for symptomatic individuals who presented with a fever, while the black data points show the same probability for individuals who did not list fever as a symptom. Plot (d) considers the respiratory rate measured for asymptomatic individuals. The plot shows the probability for *N*_*_ = 1, as a function of window center. In all cases, the error bars represent the standard error of the mean. For plots (b), (c), and (d), we approximated the standard error of a count as the square root of the count.

## IV. DISCUSSION

In this article, we showed how to compute the respiratory rate by locating the peak of the RSA feature. We computed the power spectral density from the heart rate interbeat interval time series every 5 minutes. These individual spectra were then aggregated over a night, and the respiratory rate was estimated from the averaged power spectral density. We validated our technique with the help of nasal cannula data consisting of 52 measurements obtained from 28 participants with apnea-hypopnea index < 30. The bias (mean of the predicted rate - true rate) was found to be −0.244 min^−1^ (−1.67%) while the RMS error was 0.648 min^−1^ (4.18%). The mean absolute error was 0.460 min^−1^, and the mean absolute percentage error was 3%. The absolute value of bias is larger for low values of respiratory rate. For rates lower than 16 min^−1^, the bias is −0.41 min^−1^, while for rate ≥ 16 min^−1^, the bias is 0.

We then collected respiratory rate data for 10,000 participants, ranging in age from 20−69 years, for both male and female participants. Respiratory rates measured in deep sleep (or light sleep when deep sleep data was unavailable) for adults commonly ranges from 11.8 min^−1^ - 19.2 min^−1^ (90% range). For both males and females, respiratory rate values are inversely correlated with age. From ages 20 yr - 50 yr, the Pearson *r* correlation coefficient for female (male) participants was found to be − 0.145(−0.104), while for ages > 50 yr, the corresponding values for females (males) was − 0.031(−0.043). The coefficient of variation on the other hand, increases with age (Fig.4(b)). The coefficient of variation is higher in males compared to females, for ages greater than 60, with no difference for age < 60 yr. From age 20-24 yr, the coefficient of variation measured over a 14 day period range for female (male) participants ranges from 2.3% − 9.2%(2.3%−9.5%) (90% range). For subjects in the age range 65 − 69 yr, the 90% ranges for female (male) participants are 2.5% − 16.8%(2.7% − 21.7%). Respiratory rate varies with BMI, reaching a minimum at a BMI of 25 kg/m^2^. It also varies with heart rate, increasing with increase in heart rate measured during non-REM sleep. We note however that BMI and heart rate are not independent of each other [36].

We see an interesting behavior in the way the respiratory rate varies with age for female and male participants (see Fig.4(a)). Female subjects have a higher respiratory rate than males for age < 50 yr, while for age > 50 yr, there is no difference between males and females. Female participants on average, have a higher heart rate than males [37], and we have shown that the respiratory rate is elevated in individuals with a higher heart rate. To determine whether the increased heart rate in females could contribute to the increased respiratory rate, we use Eq. 5 to obtain

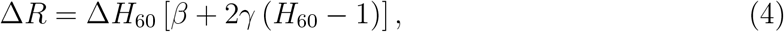

where HR is the heart rate, *H*_60_ = HR / (60 bpm), and *R* is the mean respiratory rate for individuals with a heart rate HR. For the age group 20 –24 yr, we find that male participants have ⟨*H*_60_⟩ = 1.0031. For female participants in the same age group, we find ⟨*H*_60_⟩ = 1.1123, giving us ⟨ Δ*H*_60_⟩ = 0.1092. The correlation between heart rate and respiratory rate implies that the increased heart rate can account for at most an excess of Δ*R* ≈ 0.208 min^−1^. The true difference in respiratory rates between females and males in this age group is 1.2 min^−1^ (Fig. 4(a)). The increased heart rate in females can thus account for only 17.3% of the difference between the respiratory rates of females and males. As a further test, we considered heart rate bins of 5 bpm, and selected male and female individuals within the same age bin, and the same heart rate bin. With 280 female, and 357 male participants in the heart rate bin 57.5 − 62.5 bpm, and the age bin 20 –24 yr, we find a mean respiratory rate of 16.5 min^−1^ for females, and 15.6 min^−1^ for males, with an effect size of 0.38, and a *p –*value of 1.54 × 10^−6^. Similar computations can be made for other heart rate bins and age groups. While the effect size is slightly decreased compared to the case where the heart rate is unrestricted, the increased nocturnal heart rate in females cannot solely explain the increase in respiratory rate. A striking feature seen in Fig.4(a) is the rapid decrease in the mean respiratory rate in female participants around the age ≈ 50 yr. This leads us to hypothesize that sex hormones are responsible for the difference in respiratory rates between men and women. It is well known that some sex hormones such as progesterone act as respiratory stimulants [38–40]. Since progesterone secretion decreases after menopause [38, 40], it is likely that the change in mean respiratory rate seen in females at age ≈ 50 yr is associated with menopause.

Finally, we studied how respiratory rate is affected by COVID-19. We computed respiratory rates for 3,236 uses of Fitbit devices with test dates ranging from Feb 28 - Nov 13, 2020, consisting of 2,939 symptomatic and 297 asymptomatic individuals. Let *D*_0_ be the data when symptoms first present, for symptomatic individuals, and the date when the COVID-19 test was taken, for asymptomatic individuals. We estimated the mean and standard deviation of the respiratory rate from *D*_−90_ to *D*_−30_, only considering individuals with 30 or more days of data within this date range. We obtained the mean and standard deviation for 1,247 symptomatic individuals (677 who presented with a fever, and 570 who did not) and 133 asymptomatic individuals. The *Z* –scores for each day from *D*_−28_ to *D*_+28_ are shown in Fig. 6(a) averaged over participants. For days up to *D*_−14_, the *Z* –scores are consistent with zero, but increase thereafter, reaching a maximum around ∼ *D*_+2_. The *Z* –scores decrease for larger *D*_*n*_, but interestingly, they do not fall to zero. 33% (18%) of symptomatic (asymptomatic) individuals recorded a respiratory rate 3 min^−1^ (or more) higher than their regular rate on at least one day in the 7 day window between *D*_−3_ and *D*_+3_, while only 4.6% (0.0%, limited by sample size) of symptomatic (asymptomatic) individuals showed the same increase in the 7 day period between *D*_−28_ and *D*_−22_.

In Fig. 6(b), we computed the probability of obtaining *N* ≥ *N*_*\**_ measurements satisfying *Z* ≥ 2.326. Let us estimate the noise floor by averaging the probability in the 14 day period *D*_−28_ ≤ *d* < *D*_−14_. For *N*_*\**_ = 1,3, and 5, we find noise floor values equal to 13.4%, 0.88%, and 0.092%, while the peak values are respectively, 59.3%, 23.9%, and 11.1%, yielding peak-to-noise ratios of 4.42, 27.1, and 120.4 respectively. Setting the noise floor as the false positive rate, and assuming a disease prevalence of 1 per 1000 individuals per day, we obtain positive predictive values for *N*_*\**_ = 1,3,5 to be 0.440%, 2.641%, and 10.76% respectively. For symptomatic individuals presenting with a fever (Fig. 6(c)), the *P* (*N* ≥ 1) plot peaks at 71.5%, while for symptomatic individuals who do not present with a fever, the plot peaks at 47.3%. For asymptomatic individuals (Fig. 6(d)), the plot for *N*_*\**_ = 1 peaks at 33.3%. This is smaller than for symptomatic individuals (59.3%) and for individuals who present with a fever (71.5%).

There are several limitations to the present work. The dataset of 10,000 participants consisted of individuals who were randomly selected. We did not attempt to exclude subjects with significant sleep apnea (for whom “average” respiration rate may be hard to define). Age, sex, and BMI were provided by the user, and we are unable to verify these demographic data. We have assumed that participants were healthy during the 2 week period of study, but we do not have evidence of this. This limitation is even more important for the COVID-19 study. Although we have assumed that individuals are healthy several days prior to being diagnosed with COVID-19, we do not have any way to confirm this. The date of COVID-19 diagnosis was provided by the participants themselves, and errors in this date can affect our results. Nevertheless, the results presented in this work establish that the respiratory rate is a valuable health metric which can be reliably computed using wearable devices.

## Data Availability

Fitbit's privacy policy does not permit us to make the raw data available to third parties including researchers, outside of our web API Oauth 2.0 consent process.

## ACKNOWLEDGMENTS

We thank the Fitbit users who volunteered their data for inclusion in this study. We thank the members of the Fitbit Research team for helpful discussions.

## Data availability

Fitbit’s privacy policy does not permit us to make the raw data available to third parties including researchers, outside of our web API Oauth 2.0 consent process. For specific questions, contact Fitbit at https://healthsolutions.fitbit.com/contact/.

## Author contributions

A.N. was responsible for software development, scientific analysis, and for writing the initial draft. H.W.-S. contributed to the validation of the results with observational data, and for scientific analysis. C.H. led the Fitbit COVID-19 study, contributed to project design, scientific analysis, and provided valuable insights regarding the interpretation of the data. L.B., C.O-C., and L.H. were responsible for acquiring observational data regarding respiratory rate, and for the design and execution of the data collection projects.

## Supplementary Text

Table S1 shows the details of the 2 experiments conducted to validate the respiratory rate computatons. Experiment A was conducted at Sleep Med in Columbia, SC, from Oct 17, 2019 to Nov 6, 2019. Experiment B was conducted remotely, by shipping equipment to the homes of participants, from March 9, 2020 to May 29, 2020. Participants in Experiment A wore Fitbit devices on both wrists, while participants in Experiment B wore a Fitbit device on one wrist only. We excluded participants with severe sleep apnea (Apnea-Hypopnea Index ≥30). The columns are: (i) participant ID, (ii) age, (iii) sex, (iv) experiment (A or B), (v) night of measurement (1,2, or 3), (vi) apnea-hypopnea index, (vii) wrist (left, right, or unknown), (viii) predicted respiratory rate, and (ix) true respiratory rate.

**TABLE S1:** Details of experiments A and B. The columns are: participant ID, age, sex, experiment number (A or B), night of observation (1,2, or 3), apnea hypopnea index, wrist (left, right, or unknown), predicted respiratory rate, and true respiratory rate.

| ID | Age<br>(yr) | Sex<br>(M/F) | Expt<br>(A/B) | Night<br>(1/2/3) | AHI | Wrist<br>(L/R/-) | Pred. Rate<br>(min<sup>-1</sup>) | True Rate<br>(min<sup>-1</sup>) | ID | Age<br>(yr) | Sex<br>(M/F) | Expt<br>(A/B) | Night<br>(1/2/3) | AHI | Wrist<br>(L/R/-) | Pred. Rate<br>(min<sup>-1</sup>) | True Rate<br>(min<sup>-1</sup>) |
| --- | --- | --- | --- | --- | --- | --- | --- | --- | --- | --- | --- | --- | --- | --- | --- | --- | --- |
| 1 | 35-39 | F | A | 1 | 9.2 | L | 16 | 16.4 | 13 | 45-49 | F | A | 1 | 1.5 | R | 14.4 | 14.9 |
| 1 | 35-39 | F | A | 1 | 9.2 | R | 15.6 | 16.4 | 14 | 35-39 | M | A | 1 | 2.7 | L | 14.4 | 15.2 |
| 2 | 55-59 | F | A | 1 | 17.3 | L | 15.6 | 15.9 | 14 | 35-39 | M | A | 1 | 2.7 | R | 15 | 15.2 |
| 2 | 55-59 | F | A | 1 | 17.3 | R | 15.4 | 15.9 | 15 | 45-49 | M | B | 1 | 13 | - | 13.8 | 13.8 |
| 3 | 50-54 | F | A | 1 | 1.6 | L | 12.4 | 12.4 | 15 | 45-49 | M | B | 2 | 13.2 | - | 14.2 | 14.6 |
| 3 | 50-54 | F | A | 1 | 1.6 | R | 12.4 | 12.4 | 16 | 50-54 | M | B | 2 | 4.2 | - | 18.2 | 18.1 |
| 4 | 65-69 | F | A | 1 | 5.3 | L | 13 | 13.5 | 17 | 30-34 | F | B | 1 | 25 | - | 17.8 | 16.1 |
| 4 | 65-69 | F | A | 1 | 5.3 | R | 12.8 | 13.5 | 17 | 30-34 | F | B | 2 | 12.1 | - | 16.8 | 16.6 |
| 5 | 55-59 | F | A | 1 | 10 | L | 13.8 | 14.2 | 17 | 30-34 | F | B | 3 | 14.6 | - | 17 | 17.1 |
| 5 | 55-59 | F | A | 1 | 10 | R | 14 | 14.2 | 18 | 35-39 | M | B | 1 | 4.8 | - | 13.2 | 13.6 |
| 6 | 55-59 | M | A | 1 | 26 | L | 13.4 | 13.1 | 18 | 35-39 | M | B | 2 | 5.6 | - | 13.4 | 13.7 |
| 6 | 55-59 | M | A | 1 | 26 | R | 12.8 | 13.1 | 19 | 50-54 | M | B | 1 | 7.6 | - | 17.4 | 17.5 |
| 7 | 55-59 | F | A | 1 | 7.5 | L | 20.2 | 20.2 | 19 | 50-54 | M | B | 2 | 9.8 | - | 17.4 | 17.3 |
| 7 | 55-59 | F | A | 2 | 26.3 | L | 17.4 | 17.6 | 20 | 70-74 | F | B | 1 | 18.5 | - | 15.4 | 15.2 |
| 7 | 55-59 | F | A | 2 | 26.3 | R | 16.6 | 17.6 | 21 | 45-49 | M | B | 1 | 20.1 | - | 16.4 | 16.9 |
| 8 | 50-54 | M | A | 1 | 0.6 | L | 13 | 13.6 | 21 | 45-49 | M | B | 2 | 17.2 | - | 16.8 | 16.9 |
| 8 | 50-54 | M | A | 1 | 0.6 | R | 13 | 13.6 | 22 | 50-54 | M | B | 2 | 14.4 | - | 16.2 | 16.5 |
| 9 | 50-54 | F | A | 1 | 16.9 | L | 17.4 | 17.8 | 23 | 50-54 | F | B | 1 | 4.6 | - | 12.4 | 13.3 |
| 9 | 50-54 | F | A | 1 | 16.9 | R | 17.4 | 17.8 | 23 | 50-54 | F | B | 2 | 5.8 | - | 12.8 | 13.3 |
| 10 | 45-49 | M | A | 1 | 27.1 | L | 14.2 | 14.1 | 24 | 35-39 | F | B | 1 | 14.2 | - | 17.8 | 17.4 |
| 10 | 45-49 | M | A | 1 | 27.1 | R | 14 | 14.1 | 24 | 35-39 | F | B | 2 | 12.4 | - | 18.2 | 17.7 |
| 11 | 40-44 | F | A | 1 | 2.6 | L | 15.8 | 16.1 | 25 | 40-44 | M | B | 1 | 13.8 | - | 18.2 | 16.5 |
| 11 | 40-44 | F | A | 1 | 2.6 | R | 16 | 16.1 | 26 | 50-54 | F | B | 1 | 12.5 | - | 15 | 15 |
| 12 | 40-44 | F | A | 1 | 5 | L | 13.8 | 15.3 | 26 | 50-54 | F | B | 2 | 12.6 | - | 15.6 | 15.3 |
| 12 | 40-44 | F | A | 1 | 5 | R | 13.8 | 15.3 | 27 | 45-49 | M | B | 2 | 19.3 | - | 12.6 | 12.8 |
| 13 | 45-49 | F | A | 1 | 1.5 | L | 14.6 | 14.9 | 28 | 65-69 | M | B | 1 | 6.3 | - | 13.8 | 15.7 |

## References

[1] J. A. Hirsch and B. Bishop, Respiratory sinus arrhythmia in humans: how breathing pattern modulates heart rate, Am J Physiol 241, H620 (1981).

[2] J. Hayano, F. Yasuma, A. Okada, S. Mukai, and T. Fujinami, Respiratory sinus arrhythmia. A phenomenon improving pulmonary gas exchange and circulatory efficiency, Circulation 94, 842 (1996).

[3] N. D. Giardino, R. W. Glenny, S. Borson, and L. Chan, Respiratory sinus arrhythmia is associated with efficiency of pulmonary gas exchange in healthy humans, Am J Physiol Heart Circ Physiol 284, H1585 (2003).

[4] R. R. Molgaard, P. Larsen, and S. J. Hakonsen, Effectiveness of respiratory rates in determining clinical deterioration: a systematic review protocol, JBI Database System Rev Implement Rep 14, 19 (2016).

[5] K. Mochizuki, R. Shintani, K. Mori, T. Sato, O. Sakaguchi, K. Takeshige, K. Nitta, and H. Imamura, Importance of respiratory rate for the prediction of clinical deterioration after emergency department discharge: a single-center, case-control study, Acute Med Surg 4, 172 (2017).

[6] E. Siniorakis, S. Arvanitakis, C. Tsitsimpikou, K. Tsarouhas, P. Tzevelekos, S. Panta, F. Aivalioti, C. Zampelis, F. Triposkiadis, and S. Limberi, Acute Heart Failure in the Emergency Department: Respiratory Rate as a Risk Predictor, In Vivo 32, 921 (2018).

[7] R. Strauß, S. Ewig, K. Richter, T. König, G. Heller, and T. T. Bauer, The prognostic significance of respiratory rate in patients with pneumonia: a retrospective analysis of data from 705,928 hospitalized patients in Germany from 2010-2012, Dtsch Arztebl Int 111, 503 (2014).

[8] W. S. Lim, M. M. van der Eerden, R. Laing, W. G. Boersma, N. Karalus, G. I. Town, S. A. Lewis, and J. T. Macfarlane, Defining community acquired pneumonia severity on presentation to hospital: an international derivation and validation study, Thorax 58, 377 (2003).

[9] D. Talmor, A. E. Jones, L. Rubinson, M. D. Howell, and N. I. Shapiro, Simple triage scoring system predicting death and the need for critical care resources for use during epidemics, Crit Care Med 35, 1251 (2007).

[10] J. F. Fieselmann, M. S. Hendryx, C. M. Helms, and D. S. Wakefield, Respiratory rate predicts cardiopulmonary arrest for internal medicine inpatients, J Gen Intern Med 8, 354 (1993).

[11] D. R. Goldhill, A. F. McNarry, G. Mandersloot, and A. McGinley, A physiologically-based early warning score for ward patients: the association between score and outcome, Anaesthesia 60, 547 (2005).

[12] C. P. Subbe, R. G. Davies, E. Williams, P. Rutherford, and L. Gemmell, Effect of introducing the Modified Early Warning score on clinical outcomes, cardio-pulmonary arrests and intensive care utilisation in acute medical admissions, Anaesthesia 58, 797 (2003).

[13] M. D. Howell, M. W. Donnino, D. Talmor, P. Clardy, L. Ngo, and N. I. Shapiro, Performance of severity of illness scoring systems in emergency department patients with infection, Acad Emerg Med 14, 709 (2007).

[14] D. J. Miller, J. V. Capodilupo, M. Lastella, C. Sargent, G. D. Roach, V. H. Lee, and E. R. Capodilupo, Analyzing changes in respiratory rate to predict the risk of COVID-19 infection, PLoS One 15, e0243693 (2020).

[15] A. Natarajan, H. W. Su, and C. Heneghan, Assessment of physiological signs associated with COVID-19 measured using wearable devices, NPJ Digit Med 3, 156 (2020).

[16] M. A. Cretikos, R. Bellomo, K. Hillman, J. Chen, S. Finfer, and A. Flabouris, Respiratoryrate: the neglected vital sign, Med J Aust 188, 657 (2008).

[17] J. D. Yonge, P. K. Bohan, J. J. Watson, C. R. Connelly, L. Eastes, and M. A. Schreiber, The Respiratory Rate: A Neglected Triage Tool for Pre-hospital Identification of Trauma Patients, World J Surg 42, 1321 (2018).

[18] J. Allen, Photoplethysmography and its application in clinical physiological measurement, Physiol Meas 28, 1 (2007).

[19] M. Elgendi, On the analysis of fingertip photoplethysmogram signals, Curr Cardiol Rev 8, 14 (2012).

[20] A. A. Alian and K. H. Shelley, Photoplethysmography, Best Pract Res Clin Anaesthesiol 28, 395 (2014).

[21] P. H. Charlton, D. A. Birrenkott, T. Bonnici, M. A. F. Pimentel, A. E. W. Johnson, J. Alastruey, L. Tarassenko, P. J. Watkinson, R. Beale, and D. A. Clifton, Breathing Rate Estimation From the Electrocardiogram and Photoplethysmogram: A Review, IEEE Rev Biomed Eng 11, 2 (2018).

[22] W. Karlen, S. Raman, J. M. Ansermino, and G. A. Dumont, Multiparameter respiratory rate estimation from the photoplethysmogram, IEEE Trans Biomed Eng 60, 1946 (2013).

[23] A. Schafer and K. W. Kratky, Estimation of breathing rate from respiratory sinus arrhythmia: comparison of various methods, Ann Biomed Eng 36, 476 (2008).

[24] S. Berryhill, C. J. Morton, A. Dean, A. Berryhill, N. Provencio-Dean, S. I. Patel, L. Estep, D. Combs, S. Mashaqi, L. B. Gerald, J. A. Krishnan, and S. Parthasarathy, Effect of wearables on sleep in healthy individuals: a randomized crossover trial and validation study, J Clin Sleep Med 16, 775 (2020).

[25] D. Bian, P. Mehta, and N. Selvaraj, Respiratory Rate Estimation using PPG: A Deep Learning Approach, Annu Int Conf IEEE Eng Med Biol Soc 2020, 5948 (2020).

[26] A. Natarajan, A. Pantelopoulos, H. Emir-Farinas, and P. Natarajan, Heart rate variability with photoplethysmography in 8 million individuals: a cross-sectional study, Lancet Digit Health 2, e650 (2020).

[27] C. Julien, The enigma of Mayer waves: Facts and models, Cardiovasc Res 70, 12 (2006).

[28] M. de Zambotti, F. C. Baker, A. R. Willoughby, J. G. Godino, D. Wing, K. Patrick, and M. Colrain, Measures of sleep and cardiac functioning during sleep using a multi-sensory commercially-available wristband in adolescents, Physiol Behav 158, 143 (2016).

[29] W. C. Bond, C. Bohs, J. Ebey, and S. Wolf, Rhythmic heart rate variability (sinus arrhythmia) related to stages of sleep, Cond Reflex 8, 98 (1973).

[30] J. R. Stradling, G. A. Chadwick, and A. J. Frew, Changes in ventilation and its components in normal subjects during sleep, Thorax 40, 364 (1985).

[31] D. P. White, J. V. Weil, and C. W. Zwillich, Metabolic rate and breathing during sleep, J Appl Physiol (1985) 59, 384 (1985).

[32] M. Pradella, Breathing frequency in sleep related respiratory disturbances, Arq Neuropsiquiatr 51, 227 (1993).

[33] S. Rostig, J. W. Kantelhardt, T. Penzel, W. Cassel, J. H. Peter, C. Vogelmeier, H. F. Becker, and A. Jerrentrup, Nonrandom variability of respiration during sleep in healthy humans, Sleep 28, 411 (2005).

[34] J. Krieger, N. Maglasiu, E. Sforza, and D. Kurtz, Breathing during sleep in normal middleaged subjects, Sleep 13, 143 (1990).

[35] J. Karjalainen and M. Viitasalo, Fever and cardiac rhythm, Arch Intern Med 146, 1169 (1986).

[36] G. Quer, P. Gouda, M. Galarnyk, E. J. Topol, and S. R. Steinhubl, Inter-and intraindividual variability in daily resting heart rate and its associations with age, sex, sleep, BMI, and time of year: Retrospective, longitudinal cohort study of 92,457 adults, PLoS One 15, e0227709 (2020).

[37] K. Prabhavathi, K. T. Selvi, K. N. Poornima, and A. Sarvanan, Role of biological sex in normal cardiac function and in its disease outcome - a review, J Clin Diagn Res 8, 01 (2014).

[38] A. LoMauro and A. Aliverti, Sex differences in respiratory function, Breathe (Sheff) 14, 131 (2018).

[39] A. LoMauro and A. Aliverti, Respiratory physiology of pregnancy: Physiology masterclass, Breathe (Sheff) 11, 297 (2015).

[40] M. Behan and J. M. Wenninger, Sex steroidal hormones and respiratory control, Respir Physiol Neurobiol 164, 213 (2008).

